# Rapid Tissue–CSF Water Exchange in the Human Brain Revealed by Magnetization Transfer Indirect Spin Labeling

**DOI:** 10.1101/2025.08.28.25334379

**Authors:** Yihan Wu, Kexin Wang, Licheng Ju, Anna Li, Qin Qin, Feng Xu, Doris D. M. Lin, Lawrence Kleinberg, Jiadi Xu

## Abstract

**Purpose:** To apply the Magnetization Transfer Indirect Spin Labeling (MISL) MRI technique for quantifying tissue–CSF water exchange in the human brain, and to investigate its utility in 1) visualizing perivascular spaces (PVS), and 2) characterizing altered tissue-CSF dynamics in pathologic conditions.

**Methods:** MISL was implemented on a 3T MRI using off-resonance MT to label parenchymal water and blood. The resulting exchange with CSF was captured via long-TE 3D-TSE readout to suppress parenchymal/blood signals. CSF-region-specific quantification was achieved by atlas-based segmentation. Studies were conducted in healthy subjects across age groups and in patient with metastatic brain tumor.

**Results:** MISL revealed widespread and regionally heterogeneous tissue–CSF exchange, with the strongest signals observed in the PVS and areas adjacent to the choroid plexus. MISL signals were typically 2–3% in the ventricles and subarachnoid space, and reached 3% in the cerebellar regions, suggesting tissue-to-CSF flow rates in the range of 100–300 mL/100□mL/min. The high MISL signals observed in the PVS (∼8.4%) provided enhanced sensitivity for visualizing PVS compared to T2-weighted images.

Significant age-dependent decreases in water exchange were observed across most brain regions, except for the third and fourth ventricles. In tumor patients, MISL detected elevated exchange even in areas without overt T2 hyperintensity, suggesting sensitivity to early edema formation.

**Conclusion:** MISL enables robust, non-invasive mapping of tissue–CSF exchange with high sensitivity and spatial resolution. It offers a unique window into tissue-CSF dynamics, with potential applications in PVS visualization, which reflects glymphatic function.

## Introduction

Cerebrospinal fluid (CSF) is essential to the central nervous system, playing a critical role in cushioning the brain against mechanical impact, regulating chemical homeostasis for optimal neuronal function, facilitating the removal of metabolic waste, and distributing essential nutrients (1). For over a century, the conventional understanding of CSF production centered on the choroid plexus (CP), composed of a network of capillaries and ependymal cells (2-8). It was widely believed that CSF was primarily produced through the filtration of blood plasma at the CP. However, despite decades of research, the precise mechanisms of CSF production, circulation, and clearance remain a subject of ongoing debate, particularly with the emergence of new imaging techniques and conceptual models (9-11). These findings indicate that CSF production and movement involve multiple pathways beyond the CP, with additional contributions from ependymal cells lining the ventricular walls. Moreover, the discovery of the glymphatic system has further increased the complexity of our understanding of CSF circulation (12-15). The glymphatic system suggests a more dynamic exchange between CSF and interstitial fluid (ISF). This interaction highlights the possibility that CSF circulation is not merely a passive bulk flow process but rather an active and regulated exchange influenced by physiological and pathological conditions. The uncertainty surrounding the exact mechanisms of CSF secretion and circulation has significant implications for understanding and managing CSF-related disorders, such as normal pressure hydrocephalus, idiopathic intracranial hypertension, and Alzheimer’s disease.

Stimulated by the discovery of the glymphatic system, a variety of non-invasive MRI techniques have been extensively developed to image CSF dynamics in the brain (16). Among these, phase-contrast MRI (17-19), diffusion-weighted MRI (20-23), intravoxel incoherent motion (24,25), spin-labeling methods (26-28), and fMRI methods (29) have been widely used to visualize bulk CSF flow, particularly within the ventricles. However, these techniques primarily focus on bulk CSF movement and provide limited insight into the water exchange processes occurring between CSF and surrounding brain tissues. CSF-tissue exchange is still mainly investigated by MRI contrast agents, such as intracranial (11,13,30-32) or intrathecal injection (33-35) of gadolinium-based contrast agents, as well as intravenous D-glucose infusion (36,37). While effective, these contrast-based MRI approaches are far from ideal for routine and repeated measurements in clinical settings due to safety concerns and logistical challenges. A major breakthrough in non-invasive CSF imaging came with the development of arterial spin labeling (ASL) MRI, the first non-invasive method capable of detecting CSF production from blood (38). ASL techniques have demonstrated that CSF is produced not only in the CP but also in the subarachnoid space (SAS) surrounding the cortex in the human brain (39). Other spin-labeling MRI methods have emerged to further characterize tissue-CSF exchange, including Magnetization Transfer Indirect Spin Labeling (MISL) (40,41) and Phase Alternate Labeling with Null Recovery (PALAN) (42). Both methods leverage the significant differences in protein concentration, diffusion, and T_1_/T_2_ relaxation times between CSF and brain parenchyma to track water exchange across compartments. Using these advanced techniques, rapid tissue-CSF water exchange in the mouse brain ventricles was discovered, findings that were later confirmed in human studies using a novel T_2_-labeling approach (43) and MISL with on-resonance magnetization transfer (MT) labeling approach, i.e., on-resonance MISL (44).

These advancements mark a significant shift in our understanding of CSF circulation and its interaction with brain tissues, opening new possibilities for investigating neurological diseases associated with CSF dysfunction. Further refinement of these non-invasive MRI techniques will be essential for translating these discoveries into clinical applications. Among these techniques, MISL has demonstrated significantly higher sensitivity (>10% water signal in mouse brain) (40,41) compared to the T_1_/T_2_- based approaches, making it a promising method for detecting CSF production non-invasively. However, despite its promise, MISL based on off-resonance magnetization transfer (MT) labeling has not yet been developed or validated for human brain imaging. In particular, the CSF in perivascular space (PVS)—which is not clearly visible in rodent models—has not been explored using either off-resonance or on-resonance MISL techniques. In this study, we aimed to optimize the off-resonance MISL technique for human brain applications and establish a quantitative framework for measuring tissue-CSF water exchange in the SAS, ventricles, and PVS. We also investigated age-related changes in tissue-CSF water exchange and assessed the utility of MISL in evaluating water exchange processes in brain tumors and tumor-associated edema.

## Methods

### Participants

A total of 11 healthy volunteers (age: 40±20 years; 5 females, 6 males) and one patient with a left peri-Sylvian arachnoid cyst participated, and the specific number of participants for each study is detailed below. Scanning procedures were conducted on a Philips MR Ingenia Elition 3.0T scanner (Philips Healthcare, Best, The Netherlands), utilizing a quadrature body transmit coil and a 32-channel receive head coil. One female patient in her late 40s with brain metastases was scanned on Philips 3.0 T Achieva, with the same parameters. Ethical approval was obtained from the Johns Hopkins Medicine Institutional Review Board (IRB), and all participants have provided their informed consent.

### MISL sequence

As illustrated in Fig. 1A, the MISL sequence selectively labels the water in brain parenchyma and blood via magnetization transfer (MT), while leaving CSF unaffected due to its negligible protein content. Labeled water molecules in the brain parenchyma/blood subsequently exchange with CSF, which is captured as a reduction in CSF signal intensity.

**Figure 1.**
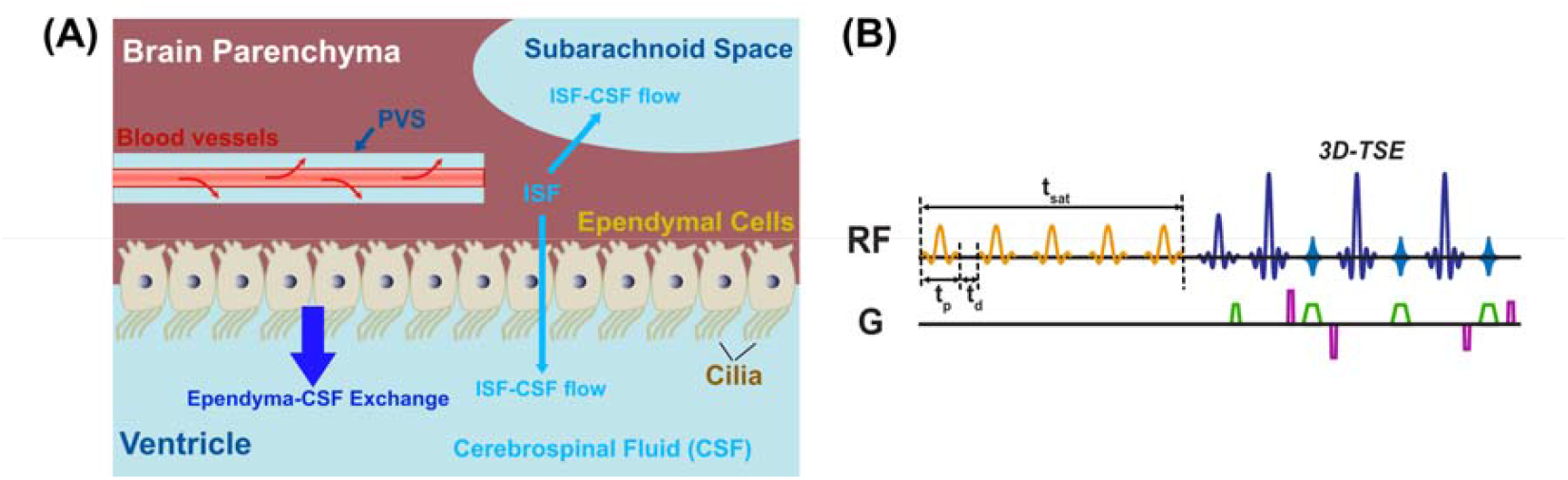
(A) Schematic illustration of the potential sources contributing to the MISL signal across the brain, including blood–CSF exchange in the perivascular space (PVS), ependymal–CSF and interstitial fluid (ISF)–CSF exchange in the ventricles, and ISF– CSF exchange in the subarachnoid space. (B) Diagram of the MISL pulse sequence. Frequency-selective magnetization transfer labeling pulses are applied to saturate water in the parenchyma or blood, followed by a long-TE 3D turbo spin-echo (3D-TSE) readout to selectively image CSF while attenuating parenchymal signal contributions.

The MISL saturation module (Fig. 1B) consists of a train of frequency-selective MT pulses, which is optimized to maximize labeling efficiency for parenchymal macromolecules while minimizing CSF direct saturation (DS). In the current study, a train of SincGaussian pulses (number of pulses: N_pulse_=50) with a peak RF amplitude of 3 μT (equivalent to 1.90 μT continuous wave), a pulse width (t_p_) of 50 ms, and an interpulse delay (t_d_) of 25 ms was used, i.e., total saturation time (t_sat_) 3.725 s. A frequency offset of −10 ppm was used for labeling unless specified, and 200 ppm for control images.

A 3D Turbo Spin-Echo (3D-TSE) readout was used for image acquisition. The MRI signal from parenchyma and blood was attenuated by a long-TE to exploit the difference in T2 relaxation times (parenchyma/blood < 100 ms vs. CSF > 1000 ms). The imaging parameters were as follows: field of view (FOV) = 160×183×150 mm^3^; slice number = 75; acquisition matrix = 80×91×75; reconstruction matrix size = 256×256×75; acquisition resolution = 2×2×2 mm^3^; TSE factor = 240 with linear ordering and startup echoes of 7; refocusing angle = 120°; compressed sensing (CS) factor = 8; echo spacing = 8.5 ms; TR/effective TE = 8 s/1081 ms. Control and label acquisitions were repeated 18 times for averaging, yielding a total scan duration of 14.5 minutes.

Labeling efficiency in the parenchyma, i.e., the parenchyma MT, was measured for each subject with identical MT saturation pulses and a single slice 2D-TSE readout (FOV = 160 × 160 × 5 mm^3^; resolution = 2 × 2 × 5 mm^3^; TSE factor = 40 with low-high ordering; refocusing angle = 120°; SENSE factor (P direction) = 2; TR/TE = 6 s / 35 ms). Two pairs of control/label images were acquired, with a scan duration of 0.5 minutes. Labeling efficiency in all tissues was assumed equivalent to the measured parenchymal MT. To quantify the water relaxation rate of parenchyma in the rotating frame 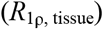, the MISL sequence was repeated using the same 2D-TSE readout parameters as in the parenchyma MT studies, while varying N_pulse_ (N_pulse_=7, 14, 27, 40, 50). As a consequence, t_sat_ =500, 1025, 2000, 2975, and 3725 ms were achieved. The mean MT signal (Δ*Z*^Tissue^) within the whole brain parenchyma in the selected slice was defined by

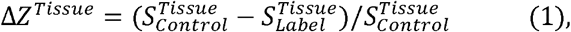

where 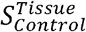 and 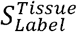 are tissue signals from the 200 ppm and -10 ppm offset acquisitions, respectively. The dependence of Δ*Z*^Tissue^ on t_sat_ was modeled using a mono-exponential recovery function to determine R_1ρ, tissue_ (45):

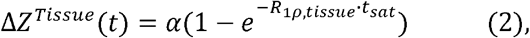

where 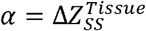 is the steady-state tissue MT signal, i.e., labeling efficiency.

### Parameter optimization

To achieve maximum labeling efficiency, full Z-spectra of the brain parenchyma were acquired by applying the same MT saturation module and the same single slice 2D-TSE readout across a range of frequency offsets (-20 to 20 ppm, step size = 1 ppm). A total of 3 subjects were included for this optimization. Four M_0_ reference images at 200 ppm were acquired and averaged for normalization. The peak saturation power (B□) for the saturation pulses was varied from 1 to 4 μT in 1 μT increments. The extent of CSF DS resulting from the MT preparation was simulated using Bloch equation modeling with only the CSF pool, assuming a T1 of 4350 ms and a T2 of 2000 ms for CSF (46). In addition, MISL sequences were acquired on three subjects using frequency offsets of ±10, ±5, ±4, ±3, -15, and -20 ppm at 3 μT peak RF amplitude, each with six pairs of control/label images to further characterize the CSF DS.

### Image processing and analysis

subtracting the label *S*_*Label*_ (offset -10 ppm) from control *S*_*Control*_ (offset 200 ppm) Similar to the tissue MT measurement in Eq.1, the MISL signal was calculated by subtracting the label *S_Label_* (offset -10 ppm) from control *S_Control_* (offset 200 ppm) images, yielding voxelwise maps of ΔZ given by

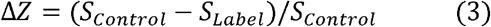

The tissue-to-CSF flow (TCF) rate (expressed in units of tissue water volume delivered per 100 unit volume of CSF per minute, mL/100 mL/min), can then be determined by: (40)

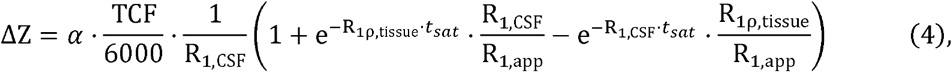

where *R*_1,app_ = *R*_1ρ,tissue_ - *R*_1,CSF_ and *R*_1*ρ*,tissue_ was measured experimentaly using Eq.2.

Three types of images can be generated from the MISL experiment: ΔS images – the difference between control and label images, calculated as Δ*S* = *S*_*control*_ - *S*_*label*_; ΔZ images – the normalized difference, calculated as ΔZ = ΔS/*S*_*control*_ and TCF maps calculated from ΔZ images using Eq. 4. In the calculation, the measured *R*_1*ρ*,tissue_ and a literature-based value for *R*_1,CSF_= 0.23s^-1^ were used (46). Both *R*_1*ρ*,tissue_ and R_1,CSF_ were assumed to be identical across all subjects, while α was individually determined for each subject as α ≈ Δ*Z*^*Tissue*^(3.725*s*).

To define regional measures of the MISL signal, T1-weighted (T1w) anatomical images from MPRAGE were acquired for image segmentation and reference space definition. The MPRAGE sequence parameters were as follows: FOV = 160×183×150 mm^3^; resolution = 1×1×1 mm^3^; TR/TE = 8.0/3.7 ms; TFE factor = 96 with linear ordering; flip angle = 8°, SENSE factor (P direction) = 2. The total scan duration of MPRAGE was 2.5 minutes. High-resolution long-TE T2w images were also acquired with TSE to generate PVS masks, with the following parameters: FOV = 160×183×150 mm^3^; resolution = 1×1×1 mm^3^; TSE factor = 240 with linear ordering and startup echoes of 57; refocusing angle = 120°; CS factor = 8; echo spacing = 8.5 ms; TR/TE = 2946 ms/1138.

Automated segmentation of gray matter, white matter, and CSF was performed in SPM12 on the T1w images. The CSF segmentation pipeline follows a similar procedure as previously described (43) and is illustrated in Fig. 2. Each subject’s T1w image was nonlinearly registered to the MNI-2009 symmetric template using the ANTs SyN framework, and deformation fields and affine transforms were inverted to propagate CerebrA atlas labels into subject space. FSL’s MCFLIRT tool was used for motion correction of the MISL images. To register MISL data, the average control and label images were rigidly aligned to the segmented CSF probability map, and the same transformation was applied to the MISL ΔZ maps to bring them into the T1w anatomical space. TCF maps were subsequently calculated from the ΔZ maps. Similarly, long-TE T2w images were also rigidly registered to the same T1w space to ensure alignment for PVS masking. A CSF mask was generated by thresholding the average control image at an empirically determined intensity cutoff to robustly isolate CSF compartments. To delineate regions expected to exhibit tissue–CSF exchange, atlas ROIs were morphologically dilated and intersected with the CSF mask. The defined ROIs are further grouped into composite categories representing the lateral ventricles (LV), third and fourth ventricles (3V and 4V), cerebral SAS, and cerebellar SAS. CP was segmented manually by the same person.

**Figure 2.**
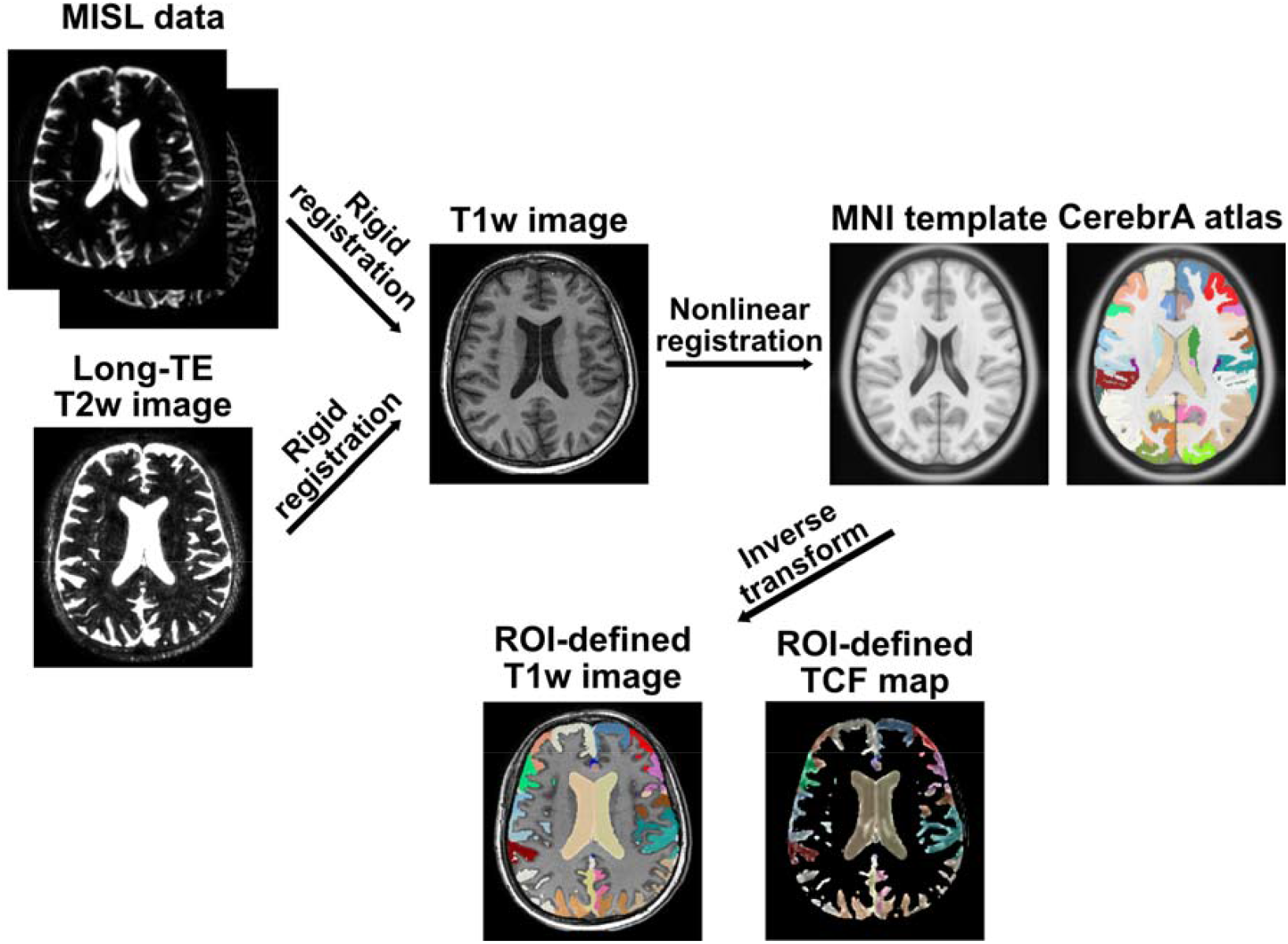
Image processing pipeline for regional tissue-CSF exchange analysis.

For the PVS mask, brain extraction was first performed on T1w images using FSL’s BET tool. The brain mask was applied to high-resolution long-TE images to exclude the extracranial signal. High-intensity voxels were then thresholded, and connected component analysis was used to remove large CSF structures. The remaining voxels were classified as PVS and applied to the ΔZ and TCF maps to extract corresponding values.

Quantitative analyses were performed to assess spatial variability and age-related trends in MISL TCF. For each subject, mean ΔZ and TCF values were calculated for each anatomically defined ROI and composite region. To assess age-related effects, linear regression models were fitted to mean TCF values within composite compartments, including the LV, 3V and 4V, cerebral SAS, cerebellar SAS, and the whole-brain CSF region. Statistical analyses were conducted using R software.

## Results

### Parameter optimization

Simulations of the CSF Z-spectrum (Fig.□3A) demonstrated that DS was negligible at a frequency offset of ±10□ppm (<□0.26%)using the MT labeling scheme applied in this study, with a peak B□ of 3□μT. Experimental Z-spectra from brain parenchyma (Fig.□3B) confirmed that MT effects increased with rising B□ amplitude, showing a sharp increase from 1 to 3□μT (MT = 10.69% to 41.59%), and only a modest additional gain at 4□μT (47.90%). Therefore, a peak B□ of 3□μT was selected for use in subsequent studies unless otherwise specified.

**Figure 3.**
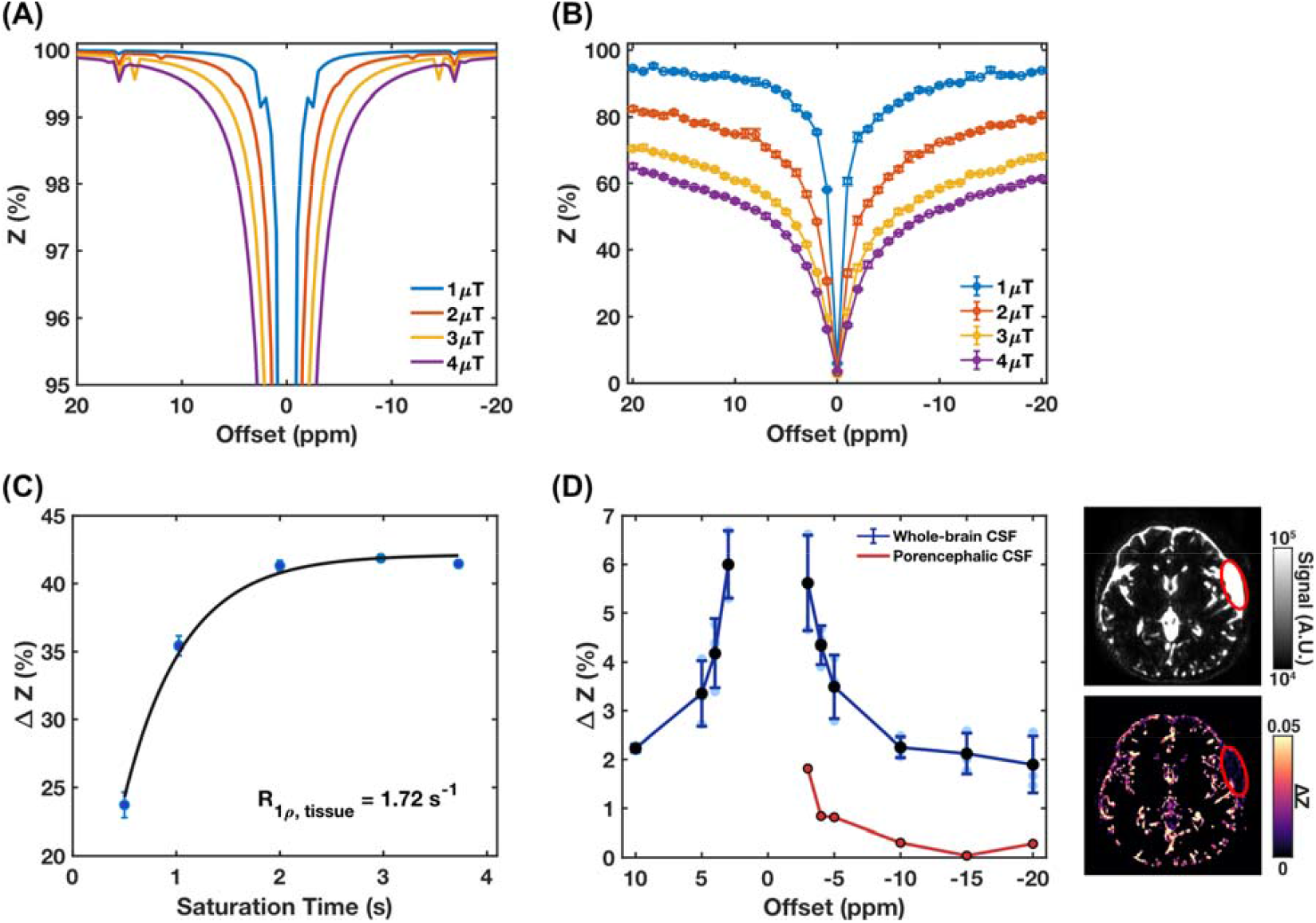
Optimization of MISL labeling parameters. (A) Simulated Z-spectrum of CSF with only water pool. (B) Experimental Z-spectra of brain parenchyma for the selected slice averaged across subjects (n=3). Saturation offsets were swept from -20 to +20□ppm in 1□ppm increments. (C) Determination of the water relaxation rate in the rotation frame 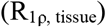. The plot shows the mean ΔZ signal within brain parenchyma (n=2) for the selected slice as a function of saturation time (500, 1025, 2000, 2975, and 3725 ms). (D) Averaged MISL ΔZ signals (n=4) from the whole-brain CSF (blue) and peri-Sylvian arachnoid cyst (red) as a function of offset. The control image and ΔZ map from the subject with a left peri-Sylvian arachnoid cyst are shown, with the peri-Sylvian arachnoid cyst indicated by a red ellipse.

The *R*_l*ρ*,tissue_ measurement results are shown in Fig.□3C, where the tissue MT signal (Δ*Z*^*Tissue*^) increased with a longer t_sat_ and reached a plateau around 2–4□s. A *R*_l*ρ*,tissue_ value of 1.72□s□^1^ was obtained through least-squares fitting using Eq.□2, with α =□0.42. Notably, the Δ*Z*^*Tissue*^ value at t_sat_ = 3.725□s (Δ*Z*^*Tissue*^(3.725s) = 0.42) was already close to α. Therefore, a = Δ*Z*^*Tissue*^(3.725s) will be used to calculate the TCF in subsequent analyses.

Whole-brain CSF ΔZ values were also evaluated across a range of frequency offsets (Fig.□3D). Within the central offsets between -5 and +5□ppm, ΔZ increased rapidly, which suggests substantial contamination from DS. In contrast, offsets beyond ±10□ppm showed a much weaker offset dependence, consistent with the expected Δ*Z*^*Tissue*^ pattern in Fig.2B and indicating minimal DS contribution—an observation in agreement with the simulation results in Fig.□2A. To experimentally quantify the DS contamination, MISL was performed on a subject with a left peri-Sylvian arachnoid cyst—a cavity filled exclusively with CSF and lacking tissue–CSF exchange. The DS signal measured in the peri-Sylvian arachnoid cyst was only 0.30% (Fig. 3), confirming that DS is negligible compared to the MISL signal (>2%) under the current MT labeling protocol. This value is also close to the simulation result (0.26%).

Based on these results, a saturation offset of -10□ppm and a peak B□ of 3□μT were selected as an optimal balance between labeling efficiency and DS suppression, making the protocol suitable for mapping tissue-CSF water exchange in regions with both fast and slow dynamics.

### Whole-brain Tissue-CSF Water Exchange Mapping

Representative MISL ΔS, ΔZ, and TCF maps from a typical healthy volunteer are shown in Fig.□4. The ΔS maps (Fig.□4A) revealed spatially heterogeneous exchange signal predominantly along the ventricular borders, cereberal SAS, cerebellar SAS, and CP, consistent with tissue-CSF water exchange occurring at these interfaces. The corresponding ΔZ maps (Fig.□4B) and TCF maps (Fig.□4C) further highlighted CSF regions adjacent to tissue—including periventricular, cereberal and cerebellar SAS, and the CP—demonstrating clear delineation with elevated exchange signal. Notably, in the zoomed-in images (Fig. 4D), a minimal ΔZ signal was observed in the central region of the LV, indicating effective suppression of CSF DS with current labeling. In contrast, strong ΔZ signals were clearly visible around the CP.

**Figure 4.**
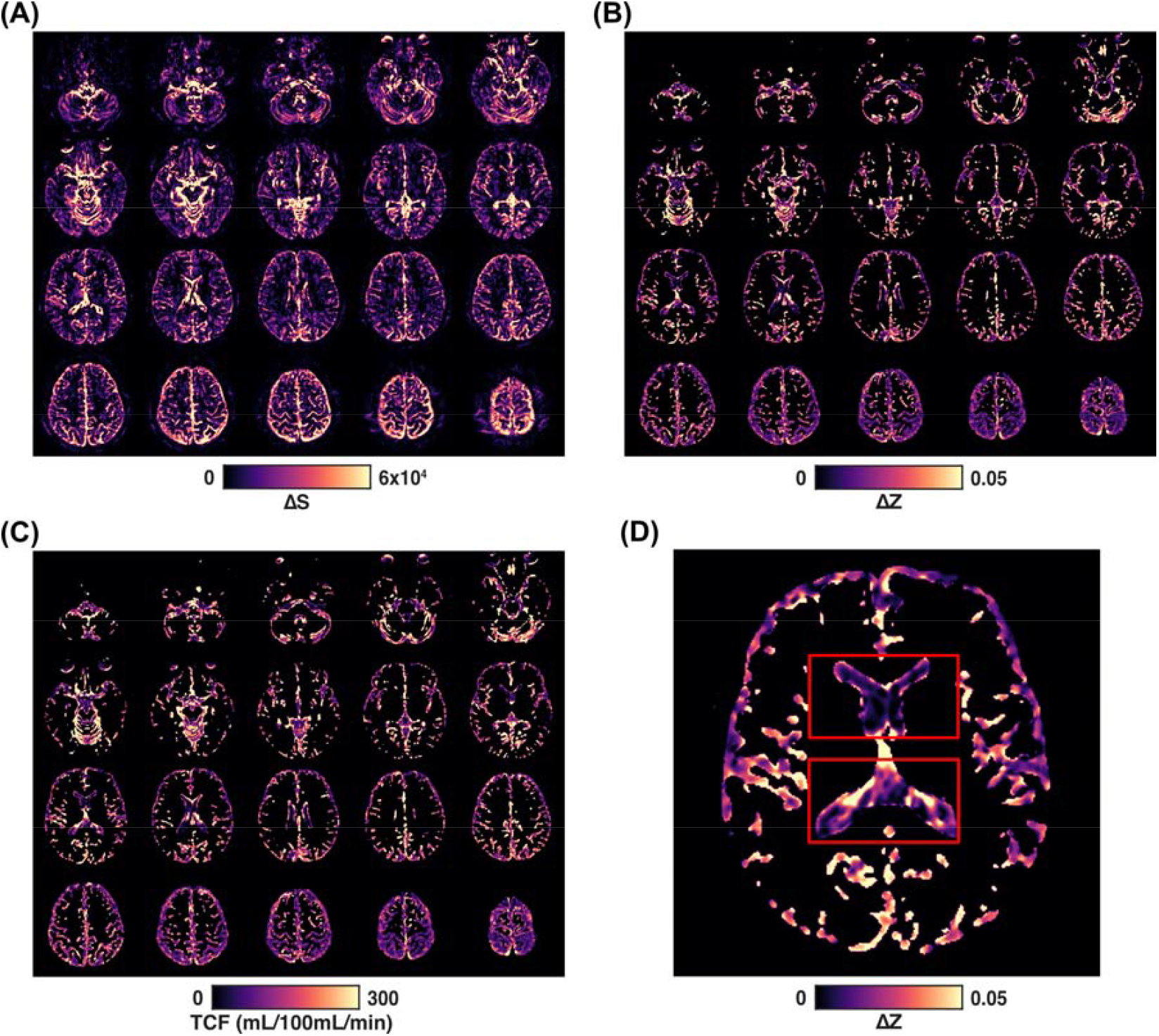
(A) Representative MISL ΔS map from a healthy volunteer, illustrating regional tissue-CSF water exchange across the brain. (B) Corresponding ΔZ maps from the same subject by applying a CSF mask, reflecting the normalized exchange-sensitive signal. (C) Corresponding TCF maps derived from the ΔZ maps for the same subject using Eq.□4. (D) Zoomed-in ΔZ map showing minimal signal in the center of the lateral ventricle and elevated signal at the ventricle–tissue interface, particularly around the choroid plexus.

### Detection of PVS with MISL

Fig.□5 shows long-TE T2w images and corresponding MISL ΔS maps acquired from two healthy subjects, one in the early 50s (Fig. 5A-C) and one in the early 20s (Fig. 5D-F). The long-TE T2w images (Fig.□5A, D) depict detailed anatomical CSF structures, including the ventricles and cortical sulci. In comparison, the MISL ΔS maps (Figs.□5B, E) provide complementary contrast sensitive to tissue–CSF water exchange and exhibit distinct signal patterns not readily visible on long-TE T2w.

**Figure 5.**
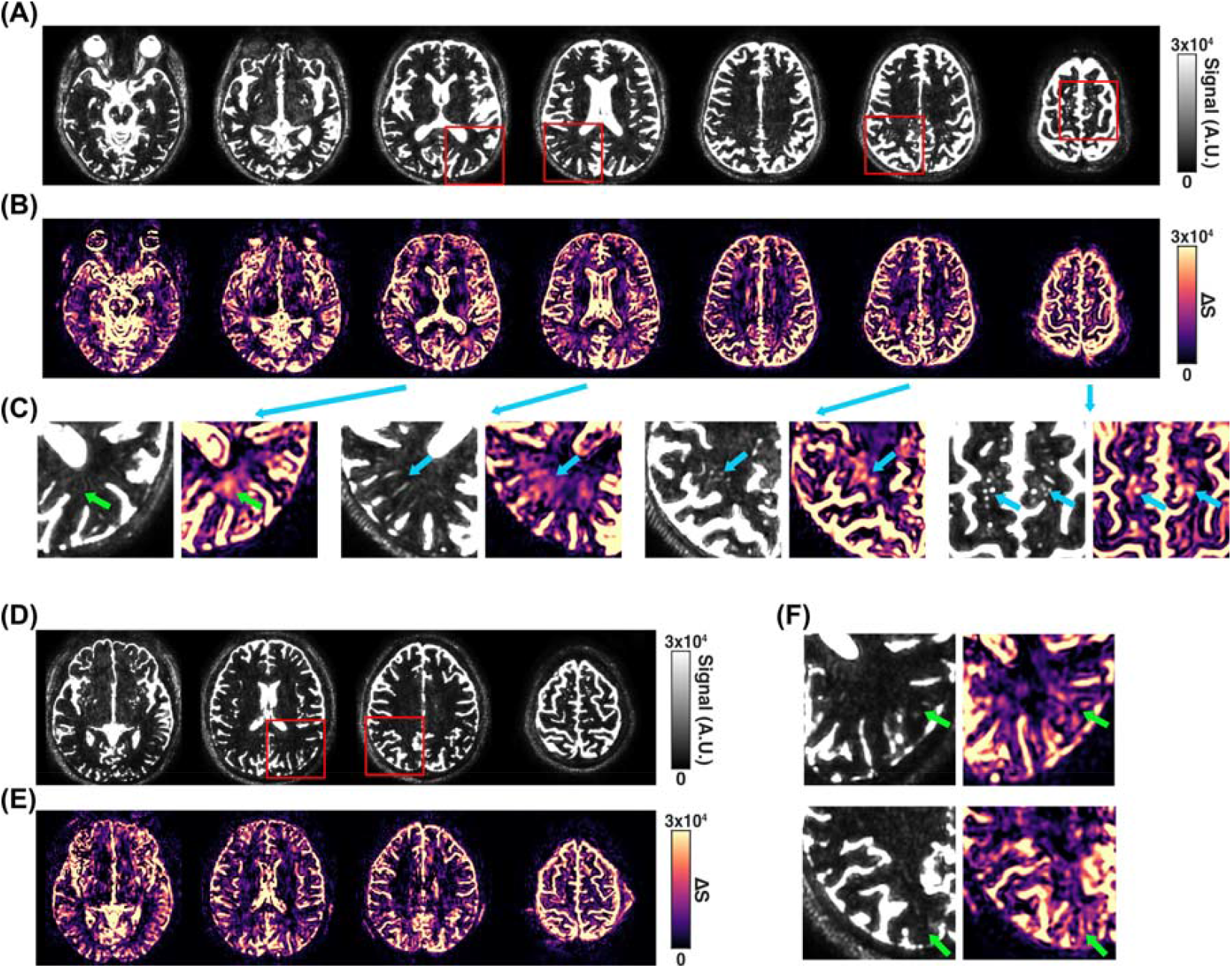
High-resolution long-TE T2w and MISL ΔS images from a healthy subject in the early 50s (A-C) and a healthy subject in the early 20s (D-F). (A, D) long-TE T2w images showing anatomical CSF distribution, including ventricles and cortical sulci. (B, E) Corresponding MISL ΔS maps revealing regional tissue-CSF water exchange. In the older subject (A–C), expanded perivascular spaces (PVS) are visible on both long-TE T2w and MISL ΔS images (C, blue arrow), while additional PVS structures are exclusively detectable in MISL maps (C, green arrow), indicating enhanced sensitivity to PVS detection by MISL. In the younger subject (C–D), PVS are largely undetectable on long-TE T2w images, but are clearly visualized with MISL as demonstrated by the zoomed images (F, green arrow).

In the older subject, zoomed-in views (Fig.□5C) highlight the locations of prominent PVSs. The most striking group of PVSs is seen in the basal ganglia, corresponding to spaces surrounding the lenticulostriate arteries. The second most noticeable PVS group appears in the centrum semiovale, following the course of medullary perforating arteries. These PVSs are clearly identifiable on both long-TE T2w images and MISL maps, confirming the high detection sensitivity of the MISL technique. Interestingly, additional PVS structures—undetectable on long-TE T2w images—are also readily visualized with MISL. This suggests that while conventional T2w imaging can capture some enlarged PVSs, MISL can detect smaller PVSs and may reveal dynamic features of PVS function not visible through static fluid contrast alone. This advantage is even more evident in younger subjects, where PVSs are typically smaller and more difficult to detect due to partial volume effects in structural imaging. Despite this, MISL successfully identifies PVSs as shown in Fig. 5F.

### Regional Variations in Tissue-CSF Water Exchange

Analysis of MISL-derived signals revealed marked spatial heterogeneity in tissue–CSF water exchange across CSF regions adjacent to cortical and subcortical structures (Fig.□6). The PVS demonstrated the highest TCF (ΔZ□=□8.42%, TCF=□550□mL/100 mL/min), followed by the CP (ΔZ□=□4.70%, TCF =□302□mL/100 mL/min), both of which are key components of fluid exchange pathways in the brain.

**Figure 6.**
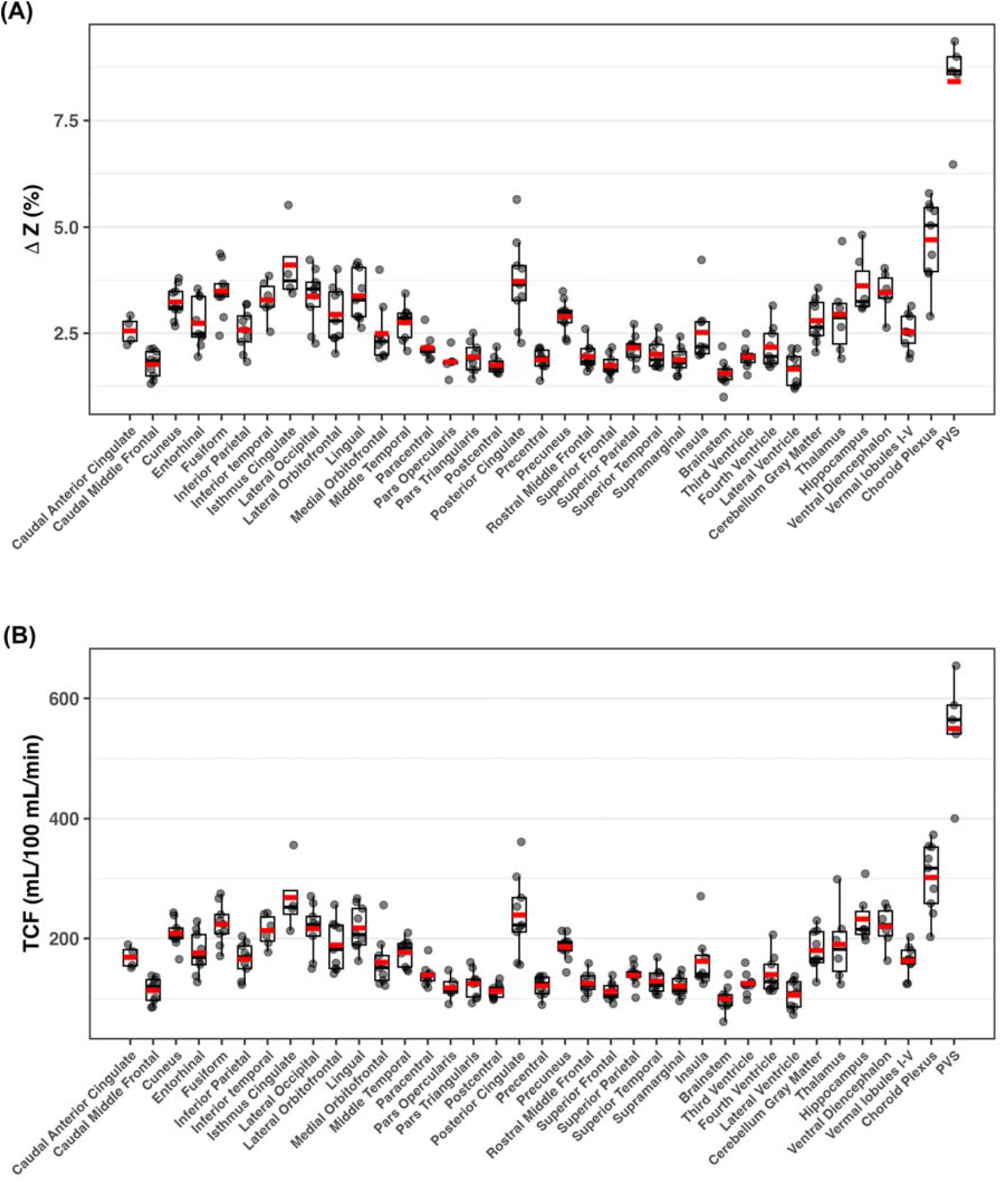
Boxplots of MISL ΔZ (A) and exchange rate (TCF) (B) across subjects (n = 9), evaluated using the CerebrA atlas, as well as perivascular space (PVS) and manually segmented regions of the choroid plexus. The strongest MISL signals are observed in the choroid plexus and PVS.

High ΔZ and TCF values were observed in CSF regions adjacent to limbic and posterior cortical areas, including the isthmus cingulate (ΔZ□=□4.11%, TCF□=□268□ mL/100 mL/min), posterior cingulate (3.72%, 239□mL/100 mL/min), and hippocampus (3.62%, 233□mL/100 mL/min). Similar elevations were found in CSF near visual and temporal cortices, such as the fusiform, lingual, lateral occipital, and inferior temporal regions, all showing ΔZ□>□3.28% and TCF□>□213□mL/100 mL/min. These patterns suggest enhanced exchange at interfaces between brain parenchyma and CSF, particularly in posterior and medial brain regions.

Subcortical-adjacent CSF regions, including those near the ventral diencephalon (ΔZ□=□3.46%, TCF□=□220□mL/100 mL/min) and thalamus (ΔZ□=2.94%, TCF□=190□mL/100 mL/min), also exhibited relatively high exchange, likely due to proximity to the ventricles and vascular structures. Adjacent CSF spaces near the precuneus, lateral orbitofrontal, entorhinal cortex, and cerebellar gray matter showed moderately elevated ΔZ (2.7–3.0%) and TCF (∼176–190□mL/100 mL/min).

In contrast, CSF regions adjacent to primary sensorimotor and frontal cortices exhibited the lowest exchange signals. These included areas near the precentral, postcentral, supramarginal, and superior frontal gyri, where ΔZ was typically <2% and TCF < 129□mL/100 mL/min. Ventricular CSF compartments such as the lateral, third, and fourth ventricles, as well as the brainstem-adjacent CSF, also demonstrated low ΔZ and TCF values.

### Age-Related Decline in Tissue-CSF Exchange Measured by MISL

We assessed the tissue-CSF exchange rate TCF across multiple CSF compartments and age groups. Linear regression analysis in Fig. 7 revealed significant age-associated decreases in TCF within the CP (slope = –2.55 mL/100 mL/min/year, p = 0.001), cerebellar SAS (–1.42 mL/100 mL/min/year, p = 0.001), whole-brain CSF (–0.75 mL/100 mL/min/year, p = 0.001), cerebral SAS (–0.63 mL/100 mL/min/year, p = 0.004), and LV (–0.91 mL/100 mL/min/year, p = 0.010). The third and fourth ventricles again showed weaker, non-significant associations with age. These findings reinforce that tissue-CSF exchange dynamics decline significantly with age, particularly in the SAS and perivascular compartments such as the CP and LV. In contrast, intraventricular spaces like the third and fourth ventricles appear less sensitive to aging-related changes in exchange.

**Figure 7.**
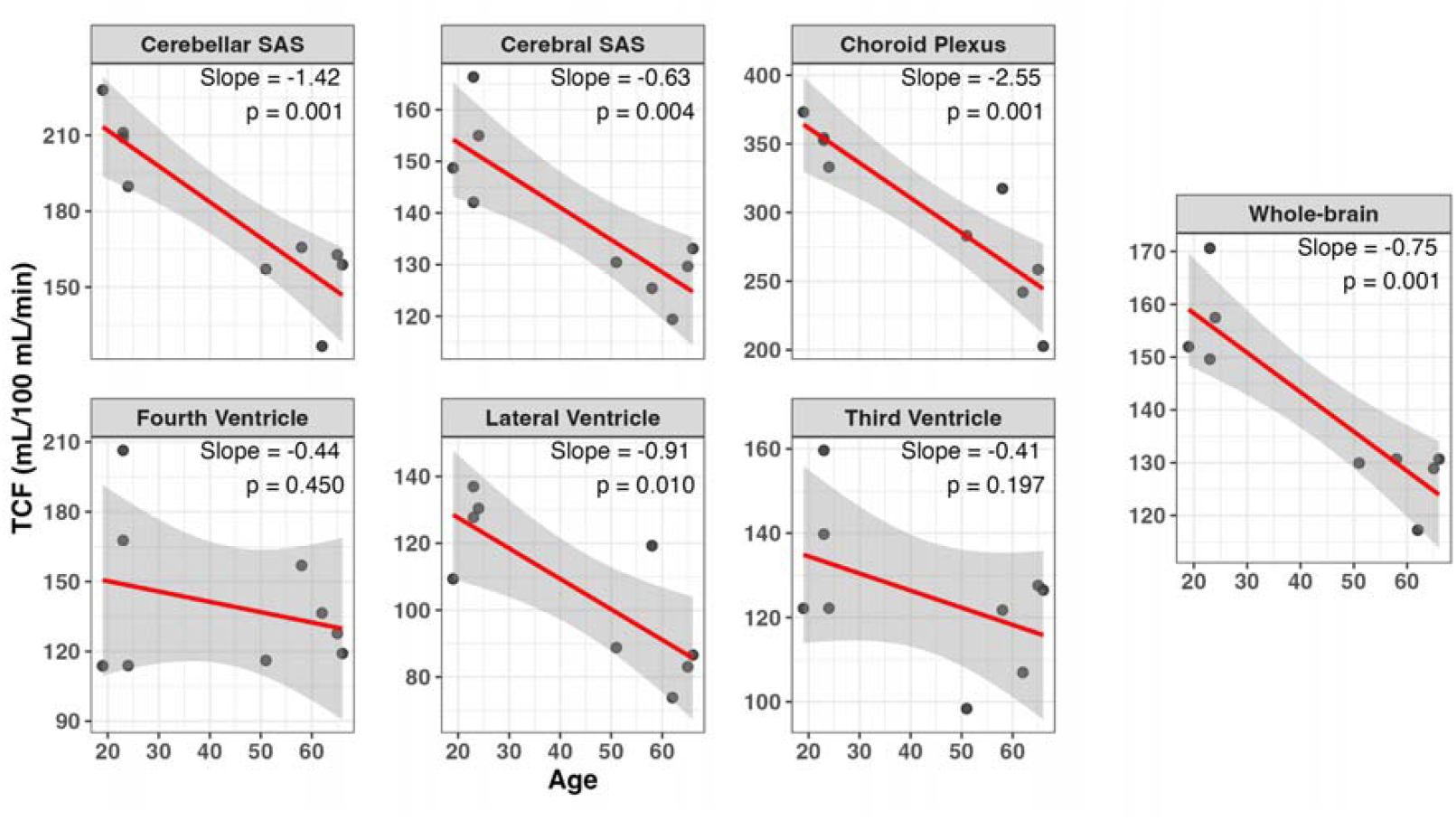
Linear regression plots show age-dependent decline in tissue-to-CSF flow rate (TCF) across various CSF compartments (n=9), including the cerebellar subarachnoid space (SAS), cerebral SAS, choroid plexus, fourth ventricle, lateral ventricles, third ventricle, and whole-brain CSF. The age-dependent slope and corresponding p-values for each region are indicated across seven CSF regions.

### Abnormal Water Exchange in Tumor and Tumor-Associated Edema

In a patient with brain metastases from primary breast cancer, MISL ΔS imaging revealed elevated water exchange signals in the tumor and adjacent tissues (Fig. 8), even when conventional FLAIR MRI did not exhibit overt hyperintensity. Zoomed-in comparisons (Fig. 8C) further demonstrated that MISL maps captured both dynamic tissue-CSF exchange (blue arrows) and structural changes, including enlarged perivascular spaces (green arrows), in regions surrounding the tumor. In areas where edema was more advanced, both MISL and T2w imaging showed abnormalities.

**Figure 8.**
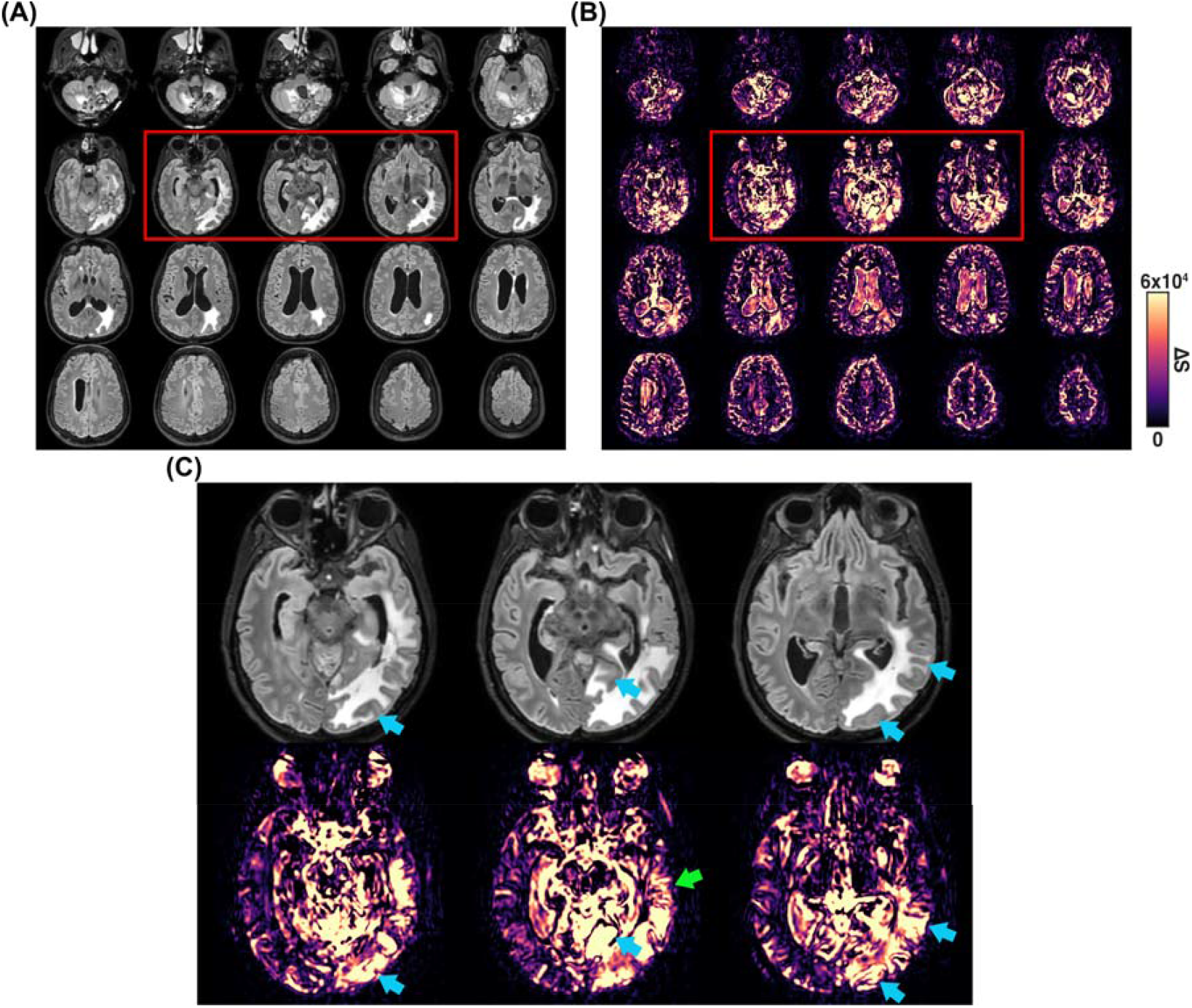
(A) FLAIR MRI of a tumor patient show edema near the tumor region. (B) MISL ΔS maps from the same tumor patient showing regional tissue-CSF water exchange. (C) Zoomed-in MISL ΔS images highlighting regions of rapid water exchange. Areas not appearing hyperintense on the FLAIR MRI but clearly detected by MISL ΔS maps are marked with blue arrows. Enlarged perivascular spaces identified by MISL are indicated with green arrows.

## Discussion

This study implements an off-resonance MISL method to detect tissue-CSF water exchange in the human brain, including the ventricles, SAS, and PVS. Simulation results and in vivo optimization demonstrated negligible contamination from DS, confirming the specificity of the method in detecting tissue-CSF water exchange. MISL revealed widespread and rapid tissue-CSF exchange across the brain, with particularly high exchange rates observed in the PVS and CP regions when comparing regional water exchange rates. We further found that tissue-CSF water exchange is strongly age-dependent, which is consistent with the animal MISL study (40) and on-resonance MISL in the human brain (44). As a proof-of-concept application, we applied MISL to assess water production in metastatic brain tumors and surrounding tissue where there is disruption of blood-brain barrier and vasogenic edema. These findings highlight the sensitivity and utility of the MISL approach for non-invasive quantification of CSF water exchange in the human brain at 3T.

When applying the MISL method for quantifying tissue-CSF water exchange, the primary confounding factor is DS. Therefore, the saturation offset, B□, and duration must be carefully optimized to minimize DS contamination. In on-resonance MT labeling, parenchymal labeling efficiency is higher than in off-resonance MT (47,48), resulting in a stronger MISL signal—typically around 3-5%, compared to ∼2-4% with off-resonance MISL. However, the major drawback of on-resonance MT is that DS is difficult to eliminate. This is particularly evident in slow-exchange regions such as the LV, where high signals persist in human brain studies using on-resonance MISL (44). Such contamination makes it less suitable for detecting slow CSF exchange dynamics. Off-resonance MISL addresses this limitation by using saturation pulses at frequency offsets well away from water, thereby greatly reducing DS. Previous studies have suggested that a short interpulse delay does not reduce labeling efficiency for slow-exchanging protons such as those involved in MT, since these labeled protons require additional time to exchange with water (49). Therefore, a 25 ms delay was used in the current study. This approach not only further minimizes DS effects but also reduces hardware demands, making it more suitable for clinical scanners. Simulations in pure water and in vivo validation using a subject with a left peri-Sylvian arachnoid cyst under the current acquisition parameters demonstrate that DS is reduced to approximately 0.3%, which is about 10% of the typical MISL signal (∼2-4%), confirming its negligible contribution. In the present study, a subject with a left peri-Sylvian arachnoid cyst was used specifically for validation. Alternatively, DS contamination can also be estimated in patients with hydrocephalus or in elderly subjects by examining regions within enlarged ventricles that are distant from the ependymal surface. Currently, we use a long-TE TSE readout for MISL. Although GRASE offers lower SAR and faster acquisition, the high acceleration (CS factor of 8) in our protocol significantly shortens the TSE readout train, making long-TE TSE both feasible and efficient for CSF exchange mapping. Moreover, it provides superior image quality compared to GRASE.

Most MISL signals observed in the SAS range from approximately 2–4%. While these values are slightly lower than those achieved with on-resonance MISL (typically 3– 5%), they are more than double the signal obtained using T2-based labeling methods (0.5–2%)□(43). This improved sensitivity enables high-resolution water exchange mapping at 2□mm resolution, even at the single-subject level. These findings align with prior animal studies, where MISL signal intensity was more than three times higher than that obtained using T1 or ADC-based labeling approaches (40,42). The measured TCF in humans is approximately ∼180□mL/100 mL/min, whereas in mice it is around 800□mL/100 mL/min—roughly 4.4 times higher (40). Interestingly, this ratio is comparable to that of cerebral blood flow (∼4), which is ∼50□mL/100 mL/min in humans□(50) versus ∼200□mL/100 mL/min in mice□(51,52). Note that MISL ΔS and ΔZ maps are highly dependent on scanner hardware and labeling efficiency, making cross-study comparisons challenging. In contrast, the TCF metric is theoretically independent of these factors, providing a more robust basis for comparison.

Significant regional variations in TCF were observed across the brain (Fig.□6), aligning with patterns previously reported using T2-based labeling methods (43). Notably, rapid water exchange was also detected in the PVS, where TCF values were substantially higher (∼8.4%) compared to those in the ventricles and SAS (2-4%). Direct visualization of the PVS is often challenging due to its small diameter and the overwhelming signal from high-intensity CSF in the ventricles and SAS. However, MISL maps successfully suppress the dominant ventricular and SAS signals, allowing the relatively weaker PVS signals to emerge more clearly. Although partial volume effects still cause the MISL ΔS signals in the PVS to appear weaker than those in larger CSF spaces, the enhancement remains significant. As shown in Fig.□5, long-TE T2w images can only identify the expanded PVS structures, but MISL provides clearer visualization of both water exchange dynamics and anatomically subtle PVSs across age groups (Fig.□5C, F). Given that the PVS plays a critical role in the glymphatic system by mediating fluid transport from the SAS to the ISF, the ability of MISL to non-invasively visualize the PVS offers a promising tool for assessing glymphatic function.

The potential sources of the MISL signal are illustrated in Fig. 1A. Due to the high water exchange rates measured, it was initially believed that the primary contributor to the MISL signal was CSF exchange with the ependymal layer, particularly in regions densely lined with epithelial cells such as the CP (40). This structure, with its extensive surface area and active transport function, consistently shows the strongest MISL signals. However, animal studies using AQP4 inhibitors have revealed that ISF to CSF exchange also contributes significantly to the MISL signal (41). This finding is further supported by results from selectively labeling ISF (44). Blood, which exhibits strong MT effects□(53), can be effectively labeled by MISL, suggesting that the MISL signal in the PVS likely reflects active fluid exchange from blood into the surrounding PVS. This water movement is tightly regulated by the blood–brain barrier (BBB), and abnormal increases in this exchange may indicate BBB dysfunction. In brain tumors and associated vasogenic edema, the breakdown of the BBB allows plasma water, proteins, and ions to leak first into the PVS due to its proximity to blood vessels, and subsequently into the broader extracellular space. This results in the expansion of both the PVS and extracellular space. The MISL signal likely captures both processes—water transport from blood into the PVS, as well as exchange between the expanded extracellular space and the intracellular space. It is important to note that traditional ASL methods with long-TE readouts primarily detect water transport from blood to ISF, and eventually into the ventricles or SAS (38,39). This multi-step process is time-consuming and results in significant decay of the labeled blood signal. Moreover, ASL captures only a small portion of the total tissue-to-CSF exchange, leading to very low signal levels, typically around 0.1-0.3% (38,39). In contrast, MISL directly labels both ISF and blood simultaneously—fluids that exchange directly with CSF—resulting in a significantly stronger signal.

Water circulation dysfunction in patients with brain tumors can be assessed using the MISL MRI. Compared to conventional FLAIR MRI imaging—which primarily reflects total water content—MISL offers a dynamic perspective by quantifying water exchange with tissue. Notably, in regions where FLAIR MRI images failed to demonstrate overt hyperintensity, MISL revealed elevated water exchange signals, suggesting early or ongoing edema formation that precedes detectable FLAIR MRI changes. This highlights the potential of MISL to capture the dynamic process of edema development, rather than just its end-state accumulation. In cases of extensive vasogenic edema, MISL also detected expansion of PVS, particularly surrounding metastatic lesions. The observed increase in free water within both tumor and tumor-adjacent regions is consistent with fluid transudation from compromised vasculature. MISL provided sensitive detection of abnormal water dynamics in the tumor not apparent in standard MRI. However, while these cross-sectional observations are promising, longitudinal studies are needed to confirm the predictive value of MISL-derived water exchange metrics for edema progression and tumor treatment response.

## Conclusions

This study demonstrates that MISL is a sensitive and specific technique for mapping tissue–CSF water exchange in the human brain. By selectively labeling parenchymal water and blood without direct saturation of CSF, MISL captures dynamic water transport processes across ventricular, subarachnoid, and perivascular compartments. Compared to conventional MRI, MISL offers enhanced sensitivity to early edema formation and glymphatic pathway dysfunction. The observed age-related reductions in exchange and the technique’s ability to visualize PVS and CP activity underscore its potential utility in both physiological and pathological contexts. Future studies will aim to establish normative values, validate predictive capability in longitudinal cohorts, and expand clinical translation across disease models.

## Data Availability

All data produced in the present study are available upon reasonable request to the authors

## Acknowledgments

This work was supported by P41EB031771, R01HL149742, P30AG066507 and R01AG080104. The authors thank Mr. Joseph S. Gillen, Mrs. Terri Lee Brawner, Ms. Kathleen A. Kahl, and Ms. Ivana Kusevic for experimental assistance.

## Notes

Grant support from NIH: P41EB031771, R01HL149742, P30AG066507 and R01AG080104

### Competing Interest Statement

The authors have declared no competing interest.

### Author Declarations

Ethical approval was obtained from the Johns Hopkins Medicine Institutional Review Board (IRB), and all participants have provided their informed consent.

